# Trends in neural tube defects in Scotland 2000-2021 prior to the introduction of mandatory folic acid fortification of non-wholemeal wheat flour: a population-based study

**DOI:** 10.1101/2024.11.22.24317787

**Authors:** Amir Kirolos, Rute Vieira, Clara Calvert, Emily Griffiths, Rachael Wood

## Abstract

**Objective:** To describe the total birth prevalence of neural tube defects (NTDs) in Scotland 2000-2021.

**Design:** National, population-based study.

**Setting:** Scotland, UK.

**Participants:** Babies with an NTD and pregnancy outcome of live birth, stillbirth (≥20 weeks gestation), or termination of pregnancy (any gestation), as recorded in the Scottish Linked Congenital Condition Dataset.

**Exposures:** Year, maternal age group, maternal area-level deprivation quintile.

**Main outcome measures:** Total birth prevalence of all NTDs and, separately, anencephaly and spina bifida (per 10,000 total births). The association between exposures and outcomes was investigated using Poisson regression.

**Results:** In Scotland 2000-2021, there were 1,178 babies with a recorded NTD (436 anencephaly, 577 spina bifida) and 1,203,491 total births. Total birth prevalence of NTDs was 9.8 [95% CI 9.2, 10.4] per 10,000 total births. Prevalence was lower among babies to mothers aged 30-39 years (compared to younger or older) and those from less (compared to more) deprived areas. There was no evidence of change over the study period in prevalence of NTDs (prevalence rate ratio [PRR] 1.01 [95% CI 0.997, 1.02] p=0.16), or spina bifida (PRR 0.99 [95% CI 0.98, 1.01] p=0.35). The prevalence of anencephaly showed no change 2000-2012, then increased 2013-2021 (PRR 1.12 [95% CI 1.06, 1.19] p<0.001).

**Conclusions:** Observed trends in anencephaly likely reflect increasing detection in early pregnancy. Current strategies are failing to equitably reduce NTDs in Scotland. Monitoring of future trends is needed to assess the impact of mandatory folic acid fortification.

**Summary:** *What is already known on this topic:*

- Recommendations for voluntary fortification of some foods, and for pre-conception daily supplements of folic acid, have been in place since the 1980s and 1990s respectively to support prevention of neural tube defects (NTDs).
- Most women do not take folic acid supplements before conception, and uptake is particularly low in younger mothers and those from poorer socioeconomic backgrounds.
- Legislation to mandate the fortification of non-wholemeal wheat flour with folic acid was introduced in November 2024. This provides an opportunity to improve the equitable prevention of NTDs.

*What this study adds:*

- There is no evidence that the total birth prevalence of NTDs in Scotland changed 2000-2021. Prevalence was lowest among mothers in the 30-39 year age group, and those from less deprived areas.
- There is no evidence that the total birth prevalence of spina bifida in Scotland changed 2000-2021, however the prevalence of anencephaly increased 2013-2021, likely reflecting increasing early antenatal detection of this condition through ultrasound scans.

*How this study might affect research, practice, or policy:*

- This study provides baseline information for the period before the introduction of mandatory fortification. It will be used to support the evaluation of mandatory fortification, and hence guide future policy development.

## Introduction

Neural tube defects (NTDs) (comprising anencephaly, spina bifida, and, least commonly, encephalocele) are congenital conditions arising due to incomplete closure of the neural tube during early fetal development(1). Pregnancies affected by an NTD are at increased risk of miscarriage or stillbirth. Babies born with spina bifida have lifelong disabilities that can include paraplegia, bladder and bowel dysfunction, hydrocephalus, and learning disabilities. Anencephaly is incompatible with long term survival. The chance of having an affected pregnancy increases with decreasing maternal levels of folate (vitamin B9) and there is well established evidence that taking pre-conception supplements of folic acid (a synthetic form of folate) reduces the risk of having an affected pregnancy(2–4).

There is evidence that the total birth prevalence (hereafter ‘prevalence’ for brevity) of NTDs fell in the UK between the 1970s and 1990s then stabilised, linked to dietary improvements and, from the mid-1980s onwards, the voluntary fortification of some breakfast cereals and bread with folic acid(5). Daily folic acid supplements have also been recommended for women who are planning, and in the first trimester of, pregnancy since 1992(6). However, a study in England in 1999-2012 found only a third reported taking supplements before conception(7). The uptake of supplements, and folate status, vary markedly by maternal sociodemographic factors(8,9).

Mandatory folic acid fortification of various staple foods introduced in other countries has been associated with a reduction in the prevalence of NTDs(10). Legislation to mandate the fortification of non-wholemeal wheat flour (and hence all products containing this flour) in the UK at a level of 250mcg per 100g was introduced in November 2024, for implementation by end 2026(11). Recommendations on daily folic acid supplements (and the possibility of additional voluntary fortification) remain unchanged. Modelling estimates this will lead to a 15-22% reduction in the prevalence of NTDs(12).

We aimed to examine secular trends in the prevalence of NTDs in Scotland 2000-2021. Establishing these pre-implementation trends will support evaluation of the impact of mandatory fortification and guide future policy decisions.

## Methods

Our study protocol is available at https://publichealthscotland.scot/publications/trends-in-neural-tube-defects-protocol/. We followed STROBE reporting guidelines(13).

### Data Sources

#### Scottish Linked Congenital Condition Dataset (SLiCCD)

SLiCCD includes all babies from pregnancies ending in Scotland 2000-2021 that were recorded as having a major congenital condition on a relevant source dataset(14,15). Major congenital conditions are defined as structural or chromosomal conditions meeting EUROCAT inclusion criteria(16). Babies are included in SLiCCD if the pregnancy ends in a live birth (any gestation, baby diagnosed before their first birthday), stillbirth (≥20 weeks gestation), or termination of pregnancy (any gestation). SLiCCD is derived from the following national datasets: statutory live birth, stillbirth (≥24 weeks gestation), and infant (<1 year) death registration records; statutory termination of pregnancy notifications; hospital maternity, neonatal, and paediatric (<1 year) inpatient discharge records; and MBRRACE-UK perinatal mortality surveillance records(17). Diagnostic codes on these source records are examined to identify records indicating a relevant condition. All records relating to an individual baby are then reconciled to provide personal identifiers; pregnancy end date and outcome; sociodemographic variables including maternal age, maternal deprivation category (Scottish Index of Multiple Deprivation (SIMD) quintile based on postcode of residence at the end of pregnancy(18)), and infant sex; and the diagnostic codes indicating the congenital condition(s) present(19). Each baby is allocated to one or more EUROCAT-defined condition group (e.g., Nervous system conditions), level 1 subgroup (e.g., Neural tube defects), and level 2 subgroup (e.g., Anencephaly, Encephalocele, Spina bifida) based on their diagnostic codes. Babies with only non-specific codes available may be assigned to no groups, or to a group but none of its subgroups. If a baby with an NTD has an underlying genetic condition, that is flagged by also allocating them to the Genetic conditions group.

#### Congenital Conditions and Rare Diseases Registration and Information Service for Scotland (CARDRISS) register

The CARDRISS register is a new, validated register of all babies with a major congenital condition from pregnancies ending in Scotland 2021 onwards(20). It has the same inclusion criteria as SLiCCD and uses the same source datasets, though CARDRISS uses a broader list of codes to identify babies that may have a registerable condition. Trained registration staff then access local clinical records to confirm whether a provisionally registered baby meets inclusion criteria and, if so, complete their registration record, including de novo coding of all congenital conditions present.

#### National Records of Scotland (NRS) live and stillbirths

Aggregate data on the number of registered live births (any gestation) and stillbirths (≥24 weeks gestation) were used as denominators to allow calculation of the total birth prevalence of NTDs. Aggregate data was obtained for each year broken down by maternal age group, maternal deprivation quintile, and infant sex.

### Statistical analysis

Statistical analyses were conducted using R v4.1.2. Code is available at https://github.com/Public-Health-Scotland/trends-in-ntds-public.

#### Number and total birth prevalence of NTDs

We used SLiCCD to report the total number of babies with an NTD from pregnancies ending 2000-2021 inclusive. We examined distribution by pregnancy outcome (live birth, stillbirth [≥20 weeks], termination of pregnancy). In line with EUROCAT guidelines, we calculated the total birth prevalence (‘prevalence’) of NTDs per 10,000 total (registered live and still [≥24 weeks]) births overall, and by maternal age group and deprivation. Total birth prevalence is the accepted proxy estimate of incidence of congenital conditions(21). We also calculated the live birth prevalence of NTDs as the number of live born babies with an NTD per 10,000 live births, overall and by infant sex (which is only reliably recorded for live births).

#### Time trend analysis

We calculated the unadjusted prevalence of NTDs for each year with 95% confidence intervals (CI) using Poisson (evidence of equidispersion of data) or Conway-Maxwell-Poisson (COM-Poisson, evidence of over/underdispersion) regression models, fitted with year as the exposure. We fitted two models: a) assuming a linear trend over time and b) using restricted cubic splines assuming a non-linear trend over time; and selected the best-fitting model using the Akaike Information Criteria (AIC).

If the model with a linear trend was chosen, we reported the unadjusted prevalence rate ratio (uPRR) for year (PRR for year estimates the average annual percentage change in prevalence) for the whole study period. If the model with a non-linear trend was chosen, we used visual inspection of plots to divide the study period into sequential sub-periods, each with a linear trend. We then used piecewise regression models to calculate the uPRRs for year for each sub-period.

#### Sociodemographic factors

We used Poisson or COM-Poisson models to calculate uPRRs for the association between prevalence of NTDs and, separately, maternal age and deprivation. To provide adjusted PRRs (aPRRs) for these associations, we extended each model to include year as an exposure (Models 1 and 2). Finally, we fitted a model including maternal age, deprivation, and year as exposures (Model 3). Models assumed a linear or non-linear time trend as per findings in the time trend analyses. Babies with missing data for maternal age and deprivation were dropped from models including these variables.

If the time trend analysis showed a linear trend, models 1-3 were also used to provide the aPRR for year adjusted for maternal age, deprivation, and both factors respectively. If the time trend analysis showed a non-linear trend, aPRRs for year for each sub-period were not provided as the relationship between the sociodemographic factors and prevalence may vary across the sub-periods making the aPRRs difficult to interpret.

All analyses were repeated for the two most common sub-types of NTDs (anencephaly and spina bifida) and for NTDs without an associated genetic condition (as NTDs in babies with an associated genetic condition are less preventable by folate supplementation(22)).

#### Validation of SLiCCD against CARDRISS

We used data from the crossover period in 2021 to calculate the positive predictive value (PPV) and sensitivity of recording of babies with NTDs in SLiCCD compared to CARDRISS(20). PPV was the percentage of babies with an NTD included in SLiCCD that were also recorded as having an NTD in CARDRISS. Sensitivity was calculated as the percentage of babies with an NTD included in CARDRISS that were also recorded as having an NTD in SLiCCD. Analyses were repeated for anencephaly, spina bifida, and NTDs without an associated genetic condition.

#### Power to detect a 20% reduction in the prevalence of NTDs

As CARDRISS rather than SLiCCD will be used for prospective evaluation of the impact of mandatory fortification, we calculated the prevalence of NTDs based on CARDRISS data for 2021. We assumed this prevalence and number of births would be the same for 2022-2024 to give us our pre-implementation data (CARDRISS data for 2022 onwards is not yet available). We then calculated the power we would have to detect a 20% reduction in the prevalence of NTDs(12,23) based on the cumulative number of births that would occur in the 10 years following full implementation of mandatory fortification. For this power calculation, we assumed the annual number of births for these 10 years would remain the same as 2021 throughout, and that the 20% reduction occurred in the first year and then did not change.

### Patient and Public involvement

We consulted with stakeholders including Spina Bifida Hydrocephalus Scotland to inform study design and interpretation.

## Results

### Number and total birth prevalence of NTDs

A total of 1,178 babies with an NTD and pregnancy end date 2000-2021 were included in SLiCCD, of which 436 (37%) had anencephaly, 165 (14%) encephalocele, and 577 (49%) spina bifida (Table 1). There were 1,203,491 total births in 2000-2021, giving a prevalence of NTDs of 9.8 [95% CI 9.2, 10.4] per 10,000 total births. Prevalence per 10,000 total births was 3.6 [95% CI 3.3, 4.0] for anencephaly, 1.4 [95% CI 1.2, 1.6] for encephalocele, and 4.8 [95%CI 4.4, 5.2] for spina bifida. Of the 1,178 babies with an NTD, 1,118 (95%) did not have an associated genetic condition (Supplementary Table 1). Overall, 35% (414) of the 1,178 babies with an NTD were live births. Pregnancy outcomes varied across the subtypes of NTD, with 8% (33) of babies with anencephaly and 53% (307) with spina bifida being live births.

**Table 1:** Number of babies with, and birth prevalence of, neural tube defects (NTDs), Scotland 2000-2021, by NTD sub-type and pregnancy outcome.

|  | Number (%) of babies with a recorded NTD | Number of total (*or live*) births | Total* (*or live*†) birth prevalence per 10,000 total (*live*) births (95% CI) |
| --- | --- | --- | --- |
| All NTDs | 1,178 | 1,203,491 | 9.8 (9.2, 10.4) |
| Live birth | 414 (35%) | 1,197,700 | 3.5 (3.1, 3.8) |
| Stillbirth (≥20 weeks) | 49 (4%) |  |  |
| Termination of pregnancy | 715 (61%) |  |  |
| Anencephaly | 436 | 1,203,491 | 3.6 (3.3, 4.0) |
| Live birth | 33 (8%) | 1,197,700 | 0.3 (0.2, 0.4) |
| Stillbirth (≥20 weeks) | 32 (7%) |  |  |
| Termination of pregnancy | 371 (85%) |  |  |
| Encephalocele | 165 | 1,203,491 | 1.4 (1.2, 1.6) |
| Live birth | 74 (45%) | 1,197,700 | 0.6 (0.5, 0.8) |
| Stillbirth (≥20 weeks) | 4 (2%) |  |  |
| Termination of pregnancy | 87 (53%) |  |  |
| Spina bifida | 577 | 1,203,491 | 4.8 (4.4, 5.2) |
| Live birth | 307 (53%) | 1,197,700 | 2.6 (2.3, 2.9) |
| Stillbirth (≥20 weeks) | 13 (2%) |  |  |
| Termination of pregnancy | 257 (45%) |  |  |

\*The total birth prevalence is calculated as the total number of babies with a recorded NTD divided by the total number of births (live and stillbirths)
†The live birth prevalence is calculated as the total number of liveborn babies with a recorded NTD divided by the total number of live births

The prevalence of NTDs was lower in maternal age groups 30-34 years and 35-39 years (8.6 [95% CI 7.7, 9.6] and 7.5 [95% CI 6.3, 8.7] per 10,000 total births respectively) compared to younger and older age groups (Table 2). Similar patterns were observed for anencephaly, spina bifida, and NTDs without an associated genetic condition (Supplementary Table 2). The prevalence of NTDs was lower in the two least deprived SIMD quintiles compared to other quintiles (8.6 [95% CI 7.4, 9.9] and 8.5 [95% CI 7.3, 9.9] per 10,000 total births in quintiles 4 and 5 respectively) (Table 2). Similar patterns were seen for spina bifida and NTDs without an associated genetic condition, though not for anencephaly (Supplementary Table 2). The live birth prevalence of NTDs, and of each NTD subtype, was lower in male compared to female live births (Table 2 and Supplementary Table 2).

**Table 2:** Number of babies with, and birth prevalence of, neural tube defects (NTDs), Scotland 2000-2021, by sociodemographic characteristics.

|  | Number (%) of babies (*or live births*) with a recorded NTD | Number (%) of total (*or live*) births | Total* (*or live †*) birth prevalence per 10,000 total (*live*) births (95% CI) |
| --- | --- | --- | --- |
| **Maternal age (years)** |  |  |  |
| <20 | 72 (6%) | 69,567 (6%) | 10.3 (8.1, 13.0) |
| 20-24 | 242 (21%) | 202,009 (17%) | 12.0 (10.5, 13.6) |
| 25-29 | 345 (29%) | 318,696 (26%) | 10.8 (9.7, 12.0) |
| 30-34 | 312 (26%) | 362,920 (30%) | 8.6 (7.7, 9.6) |
| 35-39 | 151 (13%) | 202,487 (17%) | 7.5 (6.3, 8.7) |
| ≥40 | 50 (4%) | 43,189 (4%) | 11.6 (8.6, 15.3) |
| Unknown<sup>‡</sup> | 6 (1%) | 4,623 (<1%) | - |
| **Maternal deprivation status** |  |  |  |
| SIMD quintile 1 - most deprived | 316 (27%) | 300,661 (25%) | 10.5 (9.4, 11.7) |
| SIMD quintile 2 | 261 (22%) | 247,983 (21%) | 10.5 (9.3, 11.9) |
| SIMD quintile 3 | 228 (19%) | 223,307 (19%) | 10.2 (8.9, 11.6) |
| SIMD quintile 4 | 192 (16%) | 223,729 (19%) | 8.6 (7.4, 9.9) |
| SIMD quintile 5 - least deprived | 175 (15%) | 205,286 (17%) | 8.5 (7.3, 9.9) |
| Unknown<sup>‡</sup> | 6 (1%) | 2,525 (<1%) | - |
| **Infant sex<sup>§</sup>** |  |  |  |
| *Male* | *203 (49%)* | *614,369 (51%)* | *3.3 (2.9, 3.8)* |
| *Female* | *211 (51%)* | *583,310 (49%)* | *3.6 (3.1, 4.1)* |
| *Unknown<sup>‡</sup>* | *0* | *21 (<1%)* | *-* |

\*The total birth prevalence is calculated as the total number of babies with a recorded NTD divided by the total number of births (live and stillbirths)
†The live birth prevalence is calculated as the total number of liveborn babies with a recorded NTD divided by the total number of live births
<sup>‡</sup>Total or livebirth prevalence was not calculated for groups with missing sociodemographic data as it would be misleading if patterns of missingness differed in the numerator and (unlinked) denominator data
<sup>§</sup>Information on infant sex is shown for live births only as sex is often unknown for babies with an NTD where the pregnancy ended in early stillbirth or termination of pregnancy

### NTD time trends and association with sociodemographic factors

Figure 1 shows the observed annual prevalence and unadjusted model estimates for NTDs, anencephaly, and spina bifida 2000-2021. No time trend (linear or non-linear) was identified in the prevalence of NTDs (uPRR for year 1.01 [95% CI 0.997, 1.02] p=0.16) (Table 3). For anencephaly, we found evidence of a non-linear time trend: there was no evidence of a change in prevalence 2000-2012 (uPRR 1.01 [95% CI 0.98, 1.04] p=0.64), then evidence of an increase 2013-2021 (uPRR 1.12 [95% CI 1.06, 1.19] p<0.001) (Supplementary Table 3). No time trend was identified in the prevalence of spina bifida (uPRR 0.99 [95% CI 0.98, 1.01] p=0.35) (Supplementary Table 4) or NTDs without an associated genetic condition (Supplementary Figure 1 and Supplementary Table 5).

**Figure 1:**
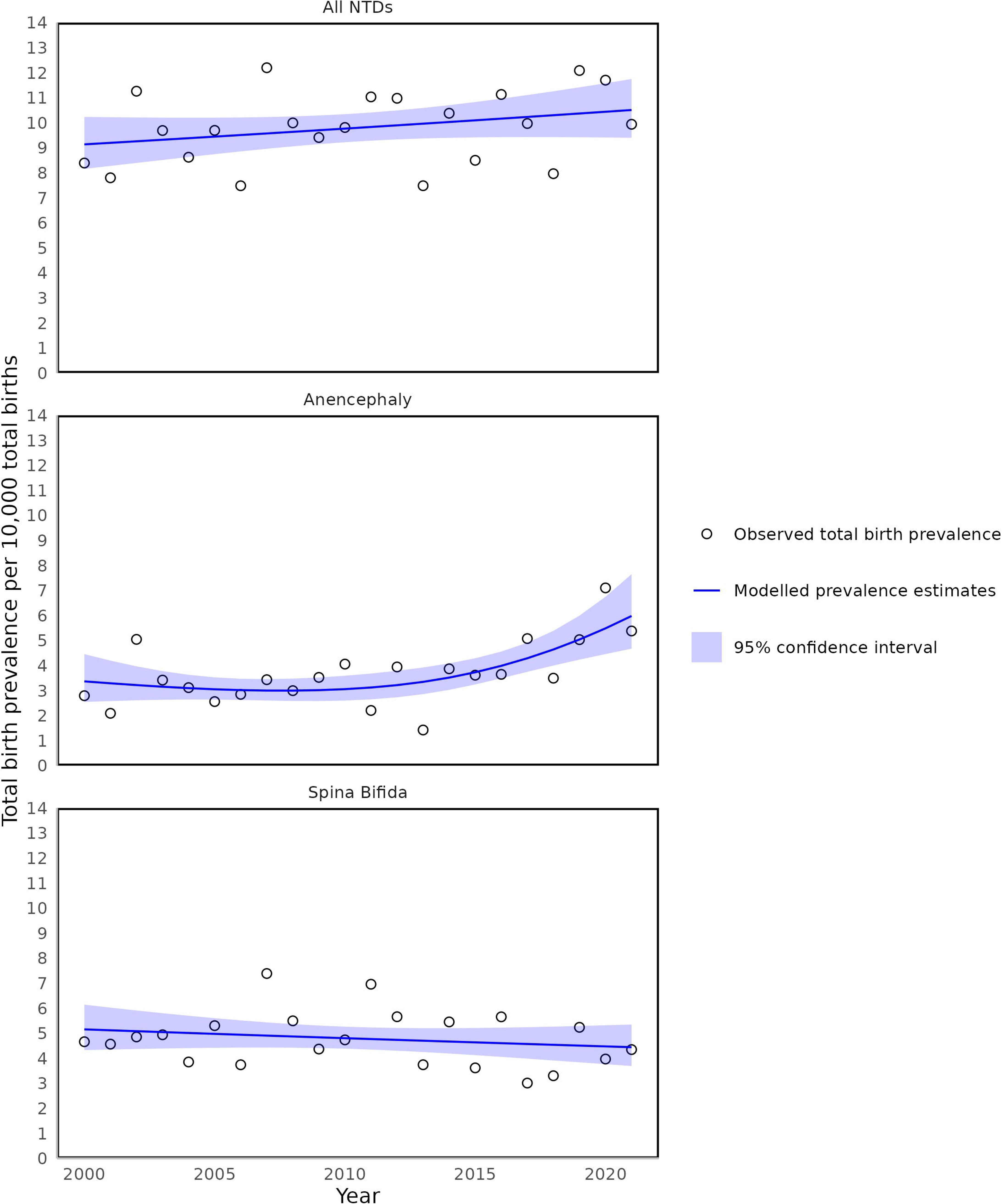
Observed and model-estimated annual total birth prevalence of all neural tube defects (NTDs), anencephaly and spina bifida, Scotland `2000-2021`

**Table 3:**
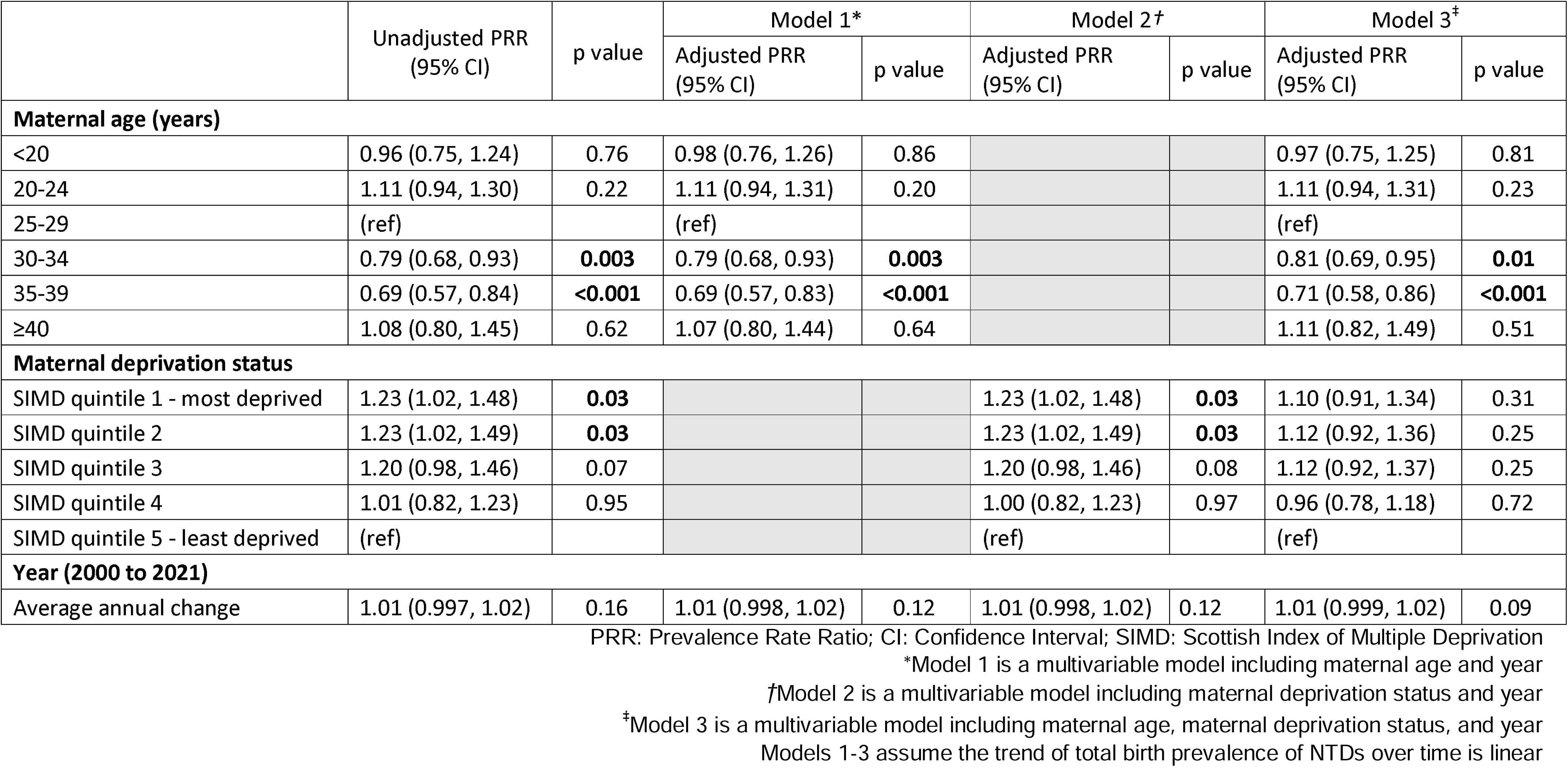
Prevalence rate ratios (PRRs) for neural tube defects (NTDs), Scotland 2000-2021.

Table 3 shows associations between maternal age and deprivation and the prevalence of NTDs. There was evidence that mothers aged 30-34 and 35-39 years old had lower aPRRs compared to those aged 25-29 years when adjusted for both maternal deprivation and year. There was no evidence of differences in aPRRs comparing each SIMD quintile to the least deprived quintile when adjusted for maternal age and year.

The time trend in the prevalence of NTDs did not change after adjusting for maternal age and deprivation in multivariable models (aPRR for year 1.01 [95% CI 0.999, 1.02] p=0.09).

### Validation of SLiCCD

Table 4 shows the number of babies with an NTD recorded in SLiCCD and/or CARDRISS in 2021. 47 of the 48 babies recorded in SLiCCD with an NTD were confirmed in CARDRISS as having an NTD, giving a PPV of 98% [95% CI 89, 100]. Of the 69 babies with an NTD recorded in CARDRISS, 47 were recorded as having an NTD in SLiCCD, giving a sensitivity of 68% [95% CI 56, 79]. Of the 22 babies that were not recorded as having an NTD in SLiCCD, 20 were included in SLiCCD in some capacity, with 12 allocated to the Nervous system condition group, but no Neural tube defect subgroup. 19 of the 22 babies had termination of pregnancy as their pregnancy outcome. PPV and sensitivity of SLiCCD compared to CARDRISS was similar for all subtypes of NTDs (Supplementary Tables 6-8).

**Table 4:** Babies recorded as having an NTD in SLiCCD and the CARDRISS register, Scotland 2021.

|  |  | CARDRISS register |  |  |
| --- | --- | --- | --- | --- |
|  |  | Recorded as NTD | Not recorded as NTD | Total |
| SLiCCD | Recorded as NTD | 47 | 1 | 48 |
|  | Not recorded as NTD | 22 |  |  |
|  | Total | 69 |  |  |

CARDRISS: Congenital conditions and rare diseases registration and information service for Scotland
SLiCCD: Scottish linked congenital condition dataset

### Power to detect a 20% reduction in the total birth prevalence of NTDs

For 2021, the prevalence of NTDs estimated from CARDRISS was 14.3 [95% CI 11.1, 18.1] per 10,000 total births. Based on this, seven years of cumulative data after full implementation of mandatory fortification will be required to reliably (i.e., with power of at least 80%) detect a 20% reduction in the prevalence of NTDs (Supplementary Table 9).

## Discussion

Our national, population-based study found no evidence of a change in the total birth prevalence of NTDs in Scotland 2000-2021. We also found no evidence of a change in the prevalence of spina bifida, or NTDs without an associated genetic condition, over this period. For anencephaly, there was no change 2000-2012, then a 12% average annual increase in prevalence 2013-2021. We found that the prevalence of NTDs was lower among babies to mothers aged 30-39 years (compared to younger or older) and to mothers from less (compared to more) deprived areas. The finding for maternal age persisted in multivariable analyses, however adjusting for maternal age and deprivation did not change our findings relating to trends over time.

Our study had strengths and limitations. We provide national, population-based data for a 22-year period with consistent policy on NTD prevention (voluntary fortification of some foods and daily folic acid supplements). We estimate the associations between NTD prevalence and maternal age and deprivation, and time trends in NTD prevalence, both unadjusted and adjusted for these key sociodemographic factors.

We used a consistent data source (SLiCCD) and validate the most recent year’s data against Scotland’s new, validated congenital condition register (CARDRISS) for transparency. The high PPV of SLiCCD compared to CARDRISS suggests we can have high confidence that babies included in our analysis were correctly categorised as having an NTD. There are known limitations in the clinical coding on source national datasets relating to terminations of pregnancy, leading to under-ascertainment of babies with an NTD and this pregnancy outcome in SLiCCD compared to CARDRISS. The total birth prevalence of NTDs based on SLiCCD data that we report is therefore likely to be an underestimate of the true prevalence. However, as there is no evidence of any change in the quality of termination data over time, the trends we observe, and our overall interpretation of results, are likely to be valid.

Our results align with a recent study assessing trends in the prevalence of NTDs in England 2000-2019(24). While this study found evidence of a small increase in the prevalence of NTDs over the study period, this was explained by an increase in anencephaly 2015-2019. We also found an increase in anencephaly over the similar period 2013-2021. Our results also align with those from an earlier study which found no change in the prevalence of NTDs without an associated genetic condition in Europe 2003-2011, using data from 28 EUROCAT registries in 19 European countries, including Wales and parts of England(25).

The prevalence of the different subtypes of NTDs would be expected to show similar trends over time(24,25), hence the recent increase in prevalence of anencephaly, but not spina bifida, seen in our results (and in England) is surprising. We cannot exclude that the increase in anencephaly is real (particularly given evidence that population folate status has worsened 2008-2019, the most recent period for which data is available(9)), or due to changes in data quality. However, given the high spontaneous loss rate of pregnancies affected by anencephaly, we suggest the most likely explanation is increasing ascertainment of affected babies in early pregnancy followed by termination of a (diagnosed) pregnancy rather than miscarriage of an (undiagnosed) pregnancy. Policy interventions from the early 2010s have promoted booking for antenatal care at earlier gestations, and a change in the method of pregnancy screening for Down’s syndrome from second trimester serum screening to first trimester combined screening, implemented in Scotland from 2011, has also promoted earlier ultrasound scanning(26,27).

We found differences in the prevalence of NTDs by sociodemographic factors, in particular maternal age. There is evidence of marked inequalities in the uptake of pre-conception folic acid supplements, with uptake particularly poor in younger women(28), and this may partly explain our findings. Mandatory fortification offers an opportunity to more effectively and equitably reduce the prevalence of NTDs, compared to the policy of voluntary fortification and supplements in place during our study period, although there is debate over whether the planned scope and level of fortification is sufficient(23,29).

Overall, this study shows that the prevalence of NTDs in Scotland 2000-2021 showed no change, and inequalities in prevalence by maternal age were evident. This study will be used to inform the evaluation of mandatory folic acid fortification of non-wholemeal wheat flour in the UK, and hence to guide future public health policy in this and other settings.

## Data availability statement

Subject to governance approvals, researchers can access pseudonymised extracts of datasets used in this study through the Scottish NHS safe haven facility supported by Public Health Scotland. Interested researchers should submit an initial enquiry form to Research Data Scotland (https://www.researchdata.scot/accessing-data/).

## Ethics Statements

### Patient consent for publication

Not applicable

### Ethics approval

This study involved the secondary analysis of routinely collected data and approval by an ethics committee was not required as informed by the NHS Health Research Authority and Public Health Scotland research risk assessment checklists. A Data Protection Impact Assessment for this study was approved by the PHS Data Protection Officer (DP22230101).

## Acknowledgements

We acknowledge the contribution of the CARDRISS analyst team within Public Health Scotland in providing relevant data extracts.

We acknowledge the contribution of our stakeholder group, including representatives from Scottish Government, Food Standards Scotland, the NHS Scotland paediatric neurosurgery service, and Spina Bifida Hydrocephalus Scotland, to study design and interpretation. The study team had final responsibility for study methodology and interpretation of results.

## Contributions

AK, RV, CC, and RW designed the study. AK, RV, CC, and EG conducted the analyses. AK drafted the manuscript, and all authors reviewed and contributed to its writing. All authors had access to the data.

## Completing interests

None declared.

