## Supplemental Material for "Trends in neural tube defects in Scotland 2000-2021 prior to the introduction of mandatory folic acid fortification of non-wholemeal wheat flour: a population-based study"

### Supplementary Tables

Supplementary Table 1: Number of babies with, and birth prevalence of, neural tube defects (NTDs) without an associated genetic condition, Scotland 2000-2021, by NTD sub-type and pregnancy outcome

|  | Number (%) of babies with a recorded NTD without a genetic condition | Number of total (*or live*) births | Total* (*or live†*) birth prevalence per 10,000 total (*live*) births (95% CI) |
| --- | --- | --- | --- |
| All NTDs | 1,118 | 1,203,491 | 9.3 (8.8, 9.9) |
| Live birth | 398 (36%) | *1,197,700* | *3.3 (3.0, 3.7)* |
| Stillbirth (≥20 weeks) | 48 (4%) |  |  |
| Termination of pregnancy | 672 (60%) |  |  |
| Anencephaly | 425 | 1,203,491 | 3.5 (3.2, 3.9) |
| Live birth | 33 (8%) | *1,197,700* | *0.3 (0.2, 0.4)* |
| Stillbirth (≥20 weeks) | 31 (7%) |  |  |
| Termination of pregnancy | 361 (85%) |  |  |
| Encephalocele | 152 | 1,203,491 | 1.3 (1.1, 1.5) |
| Live birth | 67 (44%) | *1,197,700* | *0.6 (0.4, 0.7)* |
| Stillbirth (≥20 weeks) | 4 (3%) |  |  |
| Termination of pregnancy | 81 (53%) |  |  |
| Spina bifida | 541 | 1,203,491 | 4.5 (4.1, 4.9) |
| Live birth | 298 (55%) | *1,197,700* | *2.5 (2.2, 2.8)* |
| Stillbirth (≥20 weeks) | 13 (2%) |  |  |
| Termination of pregnancy | 230 (43%) |  |  |

*The total birth prevalence is calculated as the total number of babies with a recorded NTD divided by the total number of births (live and stillbirths)

†The live birth prevalence is calculated as the total number of liveborn babies with a recorded NTD divided by the total number of live births

Supplementary Table 2: Number of babies with, and birth prevalence of, anencephaly, spina bifida and neural tube defects (NTDs) without an associated genetic condition, Scotland 2000-2021, by sociodemographic characteristics

|  | Number of total (*or* *live*) births | **Anencephaly** | | **Spina bifida** | | **NTD without an associated genetic condition** | |
| --- | --- | --- | --- | --- | --- | --- | --- |
|  |  | Number (%) of babies (*or* *live births*) | Total* (*or* *live†*) birth prevalence per 10,000 total (*live*) births (95% CI) | Number (%) of babies (*or* *live births*) | Total* (*or* *live†*) birth prevalence per 10,000 total (*live*) births (95% CI) | Number (%) of babies (*or* *live births*) | Total* (*or* *live†*) birth prevalence per 10,000 total (*live*) births (95% CI) |
| **Maternal age (years)** | | | | | | | |
| <20 | 69,567 | 27 (6%) | 3.9 (2.6, 5.6) | 35 (6%) | 5.0 (3.5, 7.0) | 69 (6%) | 9.9 (7.7, 12.6) |
| 20-24 | 202,009 | 94 (22%) | 4.7 (3.8, 5.7) | 119 (21%) | 5.9 (4.9, 7.0) | 231 (21%) | 11.4 (10.0, 13.0) |
| 25-29 | 318,696 | 122 (28%) | 3.8 (3.2, 4.6) | 170 (29%) | 5.3 (4.6, 6.2) | 326 (29%) | 10.2 (9.1, 11.4) |
| 30-34 | 362,920 | 106 (24%) | 2.9 (2.4, 3.5) | 157 (27%) | 4.3 (3.7, 5.1) | 294 (26%) | 8.1 (7.2, 9.1) |
| 35-39 | 202,487 | 69 (16%) | 3.4 (2.7, 4.3) | 70 (12%) | 3.5 (2.7, 4.4) | 145 (13%) | 7.2 (6.0, 8.4) |
| ≥40 | 43,189 | 18 (4%) | 4.2 (2.5, 6.6) | 22 (4%) | 5.1 (3.2, 7.7) | 47 (4%) | 10.9 (8.0, 14.5) |
| Unknown^‡^ | 4,623 | 0 | - | 4 (1%) | - | 6 (1%) | - |
| **Maternal deprivation status** | | | | | | | |
| SIMD quintile 1 - most deprived | 300,661 | 93 (21%) | 3.1 (2.5, 3.8) | 166 (29%) | 5.5 (4.7, 6.4) | 305 (27%) | 10.1 (9.0, 11.3) |
| SIMD quintile 2 | 247,983 | 87 (20%) | 3.5 (2.8, 4.3) | 141 (24%) | 5.7 (4.8, 6.7) | 245 (22%) | 9.9 (8.7, 11.2) |
| SIMD quintile 3 | 223,307 | 86 (20%) | 3.9 (3.1, 4.8) | 111 (19%) | 5.0 (4.1, 6.0) | 224 (20%) | 10.0 (8.8, 11.4) |
| SIMD quintile 4 | 223,729 | 93 (21%) | 4.2 (3.4, 5.1) | 72 (12%) | 3.2 (2.5, 4.1) | 178 (16%) | 8.0 (6.8, 9.2) |
| SIMD quintile 5 - least deprived | 205,286 | 75 (17%) | 3.7 (2.9, 4.6) | 83 (14%) | 4.0 (3.2, 5.0) | 160 (14%) | 7.8 (6.6, 9.1) |
| Unknown^‡^ | 2,525 | 2 (<1%) | - | 4 (1%) | - | 6 (1%) | - |
| ***Infant sex****^§^* | | | | | | | |
| *Male* | *614,369* | *10 (30%)* | *0.2 (0.1, 0.3)* | *155 (50%)* | *2.5 (2.1, 3.0)* | *196 (49%)* | *3.2 (2.8, 3.7)* |
| *Female* | *583,310* | *23 (70%)* | *0.4 (0.2, 0.6)* | *152 (50%)* | *2.6 (2.2, 3.1)* | *202 (51%)* | *3.5 (3.0, 4.0)* |
| *Unknown*^‡^ | *21* | *0* | *-* | *0* | *-* | *0* | *-* |
| *The total birth prevalence is calculated as the total number of babies with a recorded NTD divided by the total number of births (live and stillbirths)  †The live birth prevalence is calculated as the total number of liveborn babies with a recorded NTD divided by the total number of live births  ^‡^Total or livebirth prevalence was not calculated for groups with missing sociodemographic data as it would be misleading if patterns of missingness differed in the numerator and (unlinked) denominator data  §Information on infant sex is shown for live births only as sex is often unknown for babies with an NTD where the pregnancy ended in early stillbirth or termination of pregnancy | | | | | | | |

Supplementary Table 3: Prevalence rate ratios (PRRs) for anencephaly, Scotland 2000-2021

|  | Unadjusted PRR (95% CI) | p value | Model 1* | | Model 2*^†^* | | Model 3^‡^ | |
| --- | --- | --- | --- | --- | --- | --- | --- | --- |
|  |  |  | Adjusted PRR (95% CI) | p value | Adjusted PRR (95% CI) | p value | Adjusted PRR (95% CI) | p value |
| **Maternal age (years)** | | | | | | | | |
| <20 | 1.03 (0.68, 1.56) | 0.89 | 1.09 (0.72, 1.66) | 0.67 |  |  | 1.20 (0.78, 1.83) | 0.41 |
| 20-24 | 1.22 (0.93, 1.59) | 0.15 | 1.26 (0.96, 1.65) | 0.10 |  |  | 1.33 (1.01, 1.76) | **0.04** |
| 25-29 | (ref) |  | (ref) |  |  |  | (ref) |  |
| 30-34 | 0.76 (0.59, 0.99) | **0.04** | 0.75 (0.58, 0.98) | **0.03** |  |  | 0.73 (0.56, 0.95) | **0.02** |
| 35-39 | 0.89 (0.67, 1.20) | 0.45 | 0.88 (0.65, 1.18) | 0.39 |  |  | 0.83 (0.61, 1.12) | 0.22 |
| ≥40 | 1.11 (0.68, 1.82) | 0.66 | 1.07 (0.65, 1.76) | 0.79 |  |  | 1.02 (0.62, 1.68) | 0.93 |
| **Maternal deprivation status** | | | | | | | | |
| SIMD quintile 1 - most deprived | 0.85 (0.62, 1.15) | 0.28 |  |  | 0.85 (0.62, 1.17) | 0.31 | 0.72 (0.52, 0.99) | **0.04** |
| SIMD quintile 2 | 0.96 (0.71, 1.31) | 0.79 |  |  | 0.96 (0.69, 1.32) | 0.80 | 0.84 (0.61, 1.16) | 0.30 |
| SIMD quintile 3 | 1.05 (0.77, 1.43) | 0.74 |  |  | 1.06 (0.76, 1.46) | 0.74 | 0.97 (0.71, 1.33) | 0.85 |
| SIMD quintile 4 | 1.14 (0.84, 1.54) | 0.41 |  |  | 1.13 (0.82, 1.55) | 0.46 | 1.08 (0.79, 1.47) | 0.62 |
| SIMD quintile 5 - least deprived | (ref) |  |  |  | (ref) |  | (ref) |  |
| **Year**^§^ | | | | | | | | |
| Average annual change (2000-2012) | 1.01 (0.98, 1.04) | 0.64 |  |  |  |  |  |  |
| Average annual change (2013-2021) | 1.12 (1.06, 1.19) | **<0.001** |  |  |  |  |  |  |

PRR: Prevalence Rate Ratio; CI: Confidence Interval; SIMD: Scottish Index of Multiple Deprivation

*Model 1 is a multivariable model including maternal age and year

*^†^*Model 2 is a multivariable model including maternal deprivation status and year

^‡^Model 3 is a multivariable model including maternal age, maternal deprivation status, and year

Models 1-3 use restricted splines to account for the non-linear trend in total birth prevalence of anencephaly over time

^§^Due to the non-linear time trend in birth prevalence of anencephaly, we provide unadjusted estimates for the average annual change in the total birth prevalence of anencephaly for two time periods within which there was a linear trend using piecewise regression

Supplementary Table 4: Prevalence rate ratios (PRRs) for spina bifida, Scotland 2000-2021

|  | Unadjusted PRR (95% CI) | p value | Model 1* | | Model 2*^†^* | | Model 3^‡^ | |
| --- | --- | --- | --- | --- | --- | --- | --- | --- |
|  |  |  | Adjusted PRR (95% CI) | p value | Adjusted PRR (95% CI) | p value | Adjusted PRR (95% CI) | p value |
| **Maternal age (years)** | | | | | | | | |
| <20 | 0.95 (0.66, 1.37) | 0.80 | 0.93 (0.65, 1.33) | 0.69 |  |  | 0.88 (0.62, 1.27) | 0.51 |
| 20-24 | 1.11 (0.88, 1.40) | 0.40 | 1.10 (0.87, 1.39) | 0.42 |  |  | 1.06 (0.85, 1.34) | 0.60 |
| 25-29 | (ref) |  | (ref) |  |  |  | (ref) |  |
| 30-34 | 0.81 (0.65, 1.01) | 0.06 | 0.81 (0.66, 1.00) | 0.06 |  |  | 0.85 (0.69, 1.06) | 0.14 |
| 35-39 | 0.65 (0.49, 0.86) | **0.002** | 0.65 (0.49, 0.85) | **0.002** |  |  | 0.70 (0.53, 0.92) | **0.01** |
| ≥40 | 0.97 (0.63, 1.51) | 0.91 | 0.96 (0.62, 1.49) | 0.85 |  |  | 1.03 (0.66, 1.60) | 0.89 |
| **Maternal deprivation status** | | | | | | | | |
| SIMD quintile 1 - most deprived | 1.36 (1.05, 1.77) | **0.02** |  |  | 1.37 (1.08, 1.73) | **0.01** | 1.26 (0.96, 1.64) | 0.09 |
| SIMD quintile 2 | 1.40 (1.07, 1.84) | **0.01** |  |  | 1.41 (1.10, 1.80) | **0.01** | 1.31 (1.00, 1.72) | 0.05 |
| SIMD quintile 3 | 1.23 (0.92, 1.63) | 0.16 |  |  | 1.23 (0.95, 1.59) | 0.11 | 1.17 (0.88, 1.54) | 0.29 |
| SIMD quintile 4 | 0.80 (0.58, 1.09) | 0.16 |  |  | 0.80 (0.60, 1.06) | 0.12 | 0.77 (0.56, 1.05) | 0.09 |
| SIMD quintile 5 - least deprived | (ref) |  |  |  | (ref) |  | (ref) |  |
| **Year (2000-2021)** | | | | | | | | |
| Average annual change | 0.99 (0.98, 1.01) | 0.35 | 0.99 (0.98, 1.01) | 0.35 | 0.99 (0.98, 1.01) | 0.30 | 0.99 (0.98, 1.01) | 0.37 |

PRR: Prevalence Rate Ratio; CI: Confidence Interval; SIMD: Scottish Index of Multiple Deprivation

*Model 1 is a multivariable model including maternal age and year

*^†^*Model 2 is a multivariable model including maternal deprivation status and year

^‡^Model 3 is a multivariable model including maternal age, maternal deprivation status, and year

Models 1-3 assume the trend of total birth prevalence of spina bifida over time is linear

Supplementary Table 5: Prevalence rate ratios (PRRs) for neural tube defects (NTDs) without an associated genetic condition, Scotland 2000-2021

|  | Unadjusted PRR (95% CI) | p value | Model 1* | | Model 2^†^ | | Model 3‡ | |
| --- | --- | --- | --- | --- | --- | --- | --- | --- |
|  |  |  | Adjusted PRR (95% CI) | p value | Adjusted PRR (95% CI) | p value | Adjusted PRR (95% CI) | p value |
| **Maternal age (years)** | | | | | | | | |
| <20 | 0.98 (0.75, 1.26) | 0.85 | 1.00 (0.77, 1.29) | 0.97 |  |  | 0.98 (0.76, 1.27) | 0.88 |
| 20-24 | 1.12 (0.95, 1.32) | 0.19 | 1.13 (0.95, 1.33) | 0.17 |  |  | 1.12 (0.94, 1.32) | 0.21 |
| 25-29 | (ref) |  | (ref) |  |  |  | (ref) |  |
| 30-34 | 0.79 (0.68, 0.93) | **0.004** | 0.79 (0.68, 0.93) | **0.004** |  |  | 0.81 (0.69, 0.95) | **0.01** |
| 35-39 | 0.70 (0.58, 0.85) | **<0.001** | 0.70 (0.58, 0.85) | **<0.001** |  |  | 0.72 (0.59, 0.88) | **0.001** |
| ≥40 | 1.07 (0.79, 1.46) | 0.65 | 1.07 (0.79, 1.45) | 0.68 |  |  | 1.11 (0.82, 1.51) | 0.51 |
| **Maternal deprivation status** | | | | | | | | |
| SIMD quintile 1 - most deprived | 1.30 (1.07, 1.57) | **0.01** |  |  | 1.30 (1.07, 1.57) | **0.01** | 1.17 (0.96, 1.42) | 0.13 |
| SIMD quintile 2 | 1.27 (1.04, 1.54) | **0.02** |  |  | 1.26 (1.04, 1.54) | **0.02** | 1.15 (0.94, 1.41) | 0.17 |
| SIMD quintile 3 | 1.29 (1.05, 1.57) | **0.02** |  |  | 1.28 (1.05, 1.57) | **0.02** | 1.21 (0.98, 1.48) | 0.07 |
| SIMD quintile 4 | 1.02 (0.82, 1.26) | 0.85 |  |  | 1.02 (0.82, 1.26) | 0.87 | 0.98 (0.79, 1.21) | 0.83 |
| SIMD quintile 5 - least deprived | (ref) |  |  |  | (ref) |  | (ref) |  |
| **Year (2000-2021)** | | | | | | | | |
| Average annual change | 1.01 (0.998, 1.02) | 0.10 | 1.01 (0.999, 1.02) | 0.07 | 1.01 (0.999, 1.02) | 0.07 | 1.01 (0.9999, 1.02) | 0.05 |

PRR: Prevalence Rate Ratio; CI: Confidence Interval; SIMD: Scottish Index of Multiple Deprivation

*Model 1 is a multivariable model including maternal age and year

^†^Model 2 is a multivariable model including maternal deprivation status and year

‡Model 3 is a multivariable model including maternal age, maternal deprivation status, and year

Models 1-3 assume the trend of total birth prevalence of NTDs without an associated genetic condition over time is linear

Supplementary Table 6: Babies recorded as having anencephaly in SLiCCD and the CARDRISS register, Scotland 2021

|  | | CARDRISS register | | |
| --- | --- | --- | --- | --- |
|  |  | Recorded as anencephaly | Not recorded as anencephaly | Total |
| SLiCCD | Recorded as anencephaly | 26 | 0 | 26 |
|  | Not recorded as anencephaly | 11 |  |  |
|  | Total | 37 |  |  |

CARDRISS: Congenital conditions and rare diseases registration and information service for Scotland

SLiCCD: Scottish linked congenital condition dataset

Supplementary Table 7: Babies recorded as having spina bifida in SLiCCD and the CARDRISS register, Scotland 2021

|  | | CARDRISS register | | |
| --- | --- | --- | --- | --- |
|  |  | Recorded as spina bifida | Not recorded as spina bifida | Total |
| SLiCCD | Recorded as spina bifida | 20 | 1 | 21 |
|  | Not recorded as spina bifida | 7 |  |  |
|  | Total | 27 |  |  |

CARDRISS: Congenital conditions and rare diseases registration and information service for Scotland

SLiCCD: Scottish linked congenital condition dataset

Supplementary Table 8: Babies recorded as having a neural tube defect (NTD) without an associated genetic condition in SLiCCD and the CARDRISS register, Scotland 2021

|  | | CARDRISS register | | |
| --- | --- | --- | --- | --- |
|  |  | Recorded as NTD without an associated genetic condition | Not recorded as NTD without an associated genetic condition | Total |
| SLiCCD | Recorded as NTD without an associated genetic condition | 44 | 2 | 46 |
|  | Not recorded as NTD without an associated genetic condition | 23 |  |  |
|  | Total | 67 |  |  |

CARDRISS: Congenital conditions and rare diseases registration and information service for Scotland

SLiCCD: Scottish linked congenital condition dataset

Supplementary Table 9: Power to detect a 20% reduction in the total birth prevalence of neural tube defects (NTDs) based on the predicted cumulative data in the 10 years following implementation of mandatory fortification of non-wholemeal wheat flour with folic acid

| **Cumulative years following implementation of mandatory fortification** | **Total number of births*** | **Total number of births with a recorded NTD^†^** | **Total birth prevalence of NTDs per 10,000 births** | **Power to detect a 20% reduction compared with pre-implementation baseline** |
| --- | --- | --- | --- | --- |
| *Pre-implementation baseline (2021-2024)* | *193,224* | *276* | *14.3* |  |
| 1 | 48,306 | 55 | 11.4 | 0.36 |
| 2 | 96,612 | 110 | 11.4 | 0.54 |
| 3 | 144,918 | 165 | 11.4 | 0.65 |
| 4 | 193,224 | 220 | 11.4 | 0.72 |
| 5 | 241,530 | 275 | 11.4 | 0.76 |
| 6 | 289,836 | 330 | 11.4 | 0.79 |
| 7 | 338,142 | 385 | 11.4 | 0.82 |
| 8 | 386,448 | 441 | 11.4 | 0.83 |
| 9 | 434,754 | 496 | 11.4 | 0.84 |
| 10 | 483,060 | 551 | 11.4 | 0.85 |

* Assuming the total number of births in 2021 (48,306) applies for the remainder of the pre-implementation period (2022-2024) and throughout the post-implementation period

^†^ Assuming that the number of babies with an NTD recorded in the CARDRISS registry in 2021 (69) applies for the remainder of the pre-implementation period, and falls by 20% throughout the post-implementation period

The legislation introducing mandatory fortification allows a two-year transitional period, with full implementation required by end 2026. Pregnancies conceived from 2027 onwards will therefore receive its full effect.

### Supplementary Figures

Supplementary Figure 1: Observed and model-estimated annual total birth prevalence of neural tube defects (NTDs) without an associated genetic condition, Scotland 2000-2021

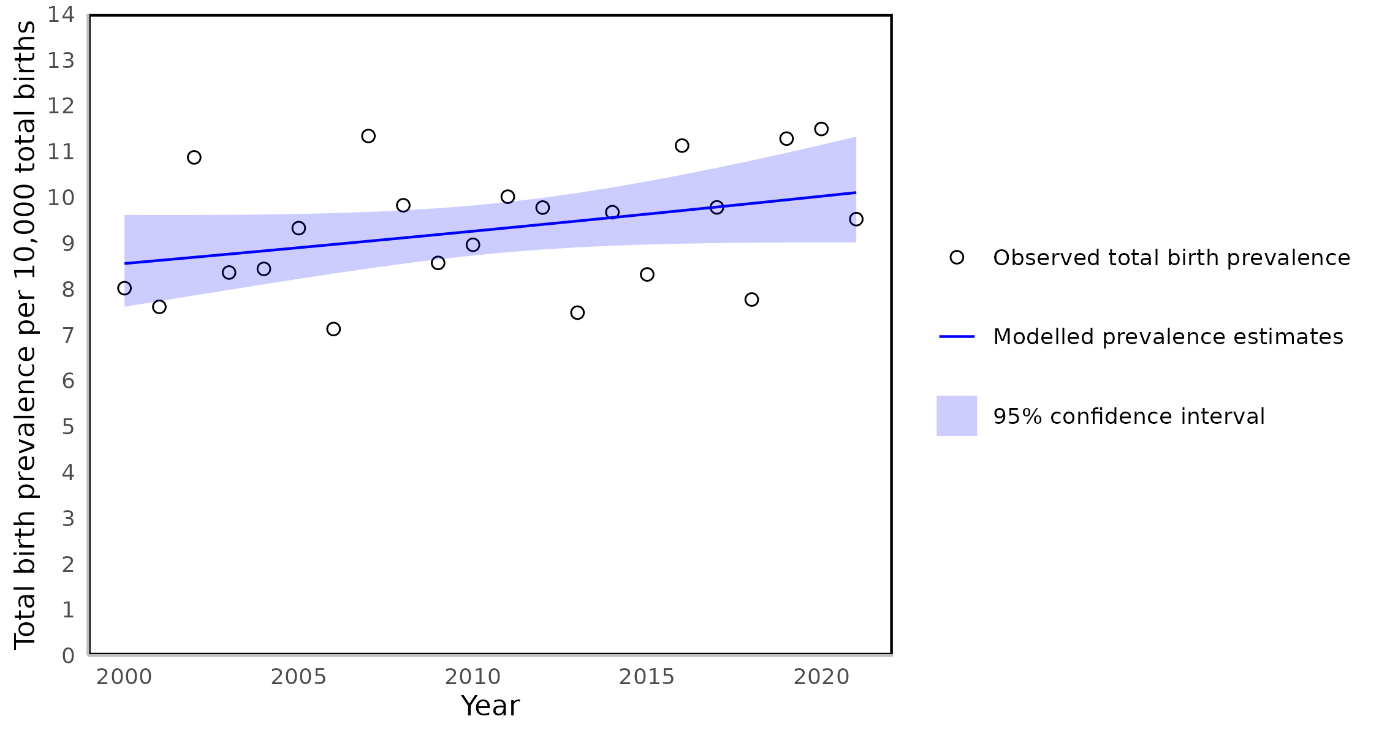

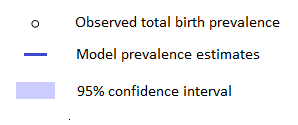
